# The first report of the prevalence of COVID-19 in Chronic myelogenous leukemia patients in the core epidemic area of China: multicentre, cross-sectional survey

**DOI:** 10.1101/2020.03.12.20034876

**Authors:** Dan-Yu Wang, Jing-Ming Guo, Zhuang-Zhi Yang, Guo-Lin Yuan, Li Meng, Wei-Ming Li

## Abstract

**Background:** Since late December 2019, the outbreak of the novel coronavirus disease, COVID-19, that began in Wuhan, has become endemic in China and more than 100 countries and regions in the world. So far, there is rare data on the prevalence of COVID-19 in patients with chronic myelogenous leukemia (CML). We aimed to describe the clinical course, outcomes of CML patients with COVID-19 and prevalence of COVID-19 in CML patients.

**Methods:** In this multicentre, cross-sectional survey, the clinical data of CML patients with COVID-19 in each center were collected. Simultaneously, an online survey was conducted for information about the CML patients under the management at each center by asking the CML patients to complete a questionnaire,from February 15, 2020 to February 21, 2020. The questionnaire includes demographic data, place of residence, smoking status, CML diagnosis and treatment, comorbidities, combined medications, epidemiological history, symptoms(fever, cough, shortness of breath, etc) during the epidemic. Additional clinical data was collected on respondents suspected or confirmed to have COVID-19. We described and analyzed the prevalence of COVID-19 in CML patients, and focus on the clinical characteristics and outcomes of COVID-19 patients. Data were compared between the CML patients with optimal response and those with non-optimal response. The primary outcome was prevalence of COVID-19 in CML patients, as of Feb 21, 2020. Secondary outcomes included the history of epidemiology of CML patients, the clinical characteristics and outcomes of CML patients with COVID-19.

**Findings:** Of 392 respondents, 223(56.9%) were males, and 240(61.2%) were 50 years or younger. Only 10 patients took drugs irregularly due to the influence of the epidemic because of traffic control, pharmacies unable to operate normally, etc. In the history of epidemiology, there were 4 patients with definite contact with COVID-19, of which 3 were remote contact and 1 was close contact. 12 respondents had fever, cough or shortness of breath during the epidemic, 1 case (common type) was confirmed with COVID-19 and cured after treatment. 1 patient was clinically diagnosed and succumbed. 1 of 299 (0.3%) patients with an optimal response was diagnosed with COVID-19. Of the 50 patients who failed to respond to CML treatment or had a poor response, 1 patient (2%) had a clinical diagnosis of COVID-19.

**Interpretation:** While the 392 CML respondents required regular referrals to hospitals, they did not have much contact with COVID-19 patients during the outbreak. Patients who failed to achieved an optimal response to CML therapy appear more likely to have a symptomatic infection with SARS-CoV-2. Older patients with comorbidities are at increased risk of death.

**Funding:** This work was supported by grants from the National Natural Science Foundation of China(NSFC)(81873440&81700142).

## Introduction

Since late December 2019, the outbreak of the 2019 novel coronavirus disease (COVID-19) in Wuhan, China, has been endemic in China and more than 100 countries and regions in the world.^1-7^ By March 1, 2020, China had reported a total of 80,026 confirmed cases of novel coronavirus disease, with 2,912 deaths. In Hubei province which is still the core area of the COVID-19 epidemic in China there were 67,103 cases, accounting for 83.9% of total Chinese cases. The number of confirmed cases in South Korea, Iran, Italy, France, Japan and other countries continues to rise.

Some studies have suggest that cancer patients are more susceptible to infection with SARS-CoV-2 than healthy people and have a worse prognosis because their immune systems are suppressed by the effects of the tumors and anti-cancer treatment ^8^. But this view is controversial ^9,10^.

Chronic myelogenous leukemia (CML) is a neoplastic disease of hematopoietic stem cells that has an annual incidence of 0.4-1.75/100,000 ^11-15^ and accounts for 15% of adult leukemia. Since imatinib and subsequent generations of newer tyrosine kinase inhibitors (TKIs) have been used with CML, the prognosis of CML patients has improved significantly. In fact, CML patients are currently approaching normal life expectancy. However, as the survival rates of CML patients improves, the prevalence of CML is increasing. The prevalence of CML in France is estimated to reach 18 / 100,000 in 2018 and 24 / 100,000 in 2030 ^16^. We sought to determine whether CML patients have been as susceptible to SARS-CoV-2 as other cancer patients during the current epidemic, and what their prognosis has been. Standardized management of this large number of patients has become a new challenge for our hematologists. In order to optimize the care of CML patients as the pandemic spreads, it is therefore essential to understand the characteristics of SARS-CoV-2 infections in CML patients and the experience of their management in Hubei province.

## Methods

### Study design and participants

When the outbreak started, the 29 centers of the Hubei Anti-Cancer Association Chronic Myeloid Leukemia Standardized Management Collaboration Group adopted comprehensive management measures for CML patients. Various kinds of online consultation, online smart hospitals and online courses were made available to provide patients with information while they remained at home. Individualized treatment schemes were proposed according to the clinical situation of each patient. It was recommended that all patients, especially those with stable conditions, refrain from leaving their homes, and even avoid going to hospitals. Volunteers were organized to deliver drugs to CML patients in their homes in order to ensure their normal treatment would not be interrupted. Patients with COVID-19-related symptoms were promptly diagnosed and treated accordingly.

All patients were diagnosed with 2019 novel coronavirus disease, according to WHO interim guidance^17^ or the 5th version of the novel coronavirus infection pneumonia diagnosis and treatment (The National health commission, People’s Republic of China)^18^. Laboratory confirmation of SARS-CoV-2 infection was performed by the local health authority. Identification of COVID-19 patients was achieved by reviewing and analysing admission logs and histories from all available electronic medical records and patient care resources. Efficacy of CML was assessed according to the 2020 ELN guidelines^19^. The investigation was approved by the ethics committee of Union hospital, Tongji Medical college, Huazhong University of science and Technology.

### Data collection

From February 15, 2020 to February 21, 2020, the clinical data of CML patients with COVID-19 in each center were collected. Simultaneously, an online survey was conducted for information about the CML patients under the management at each center by asking the CML patients to complete a questionnaire. A total of 413 questionnaires were collected, of which 392 were valid, but 21 of these were excluded because the respondents did not live in Hubei province during the outbreak. The questionnaire asked about general demographic characteristics, place of residence, smoking status, CML diagnosis and treatment, comorbidities, combined medications, epidemiological history, and symptoms of fever, cough, shortness of breath, etc. during the epidemic. Additional clinical data was collected on respondents suspected or confirmed to have COVID-19. Any missing or uncertain records were collected and clarified through direct communication with involved health-care providers and their families.

### Outcomes

The primary outcome was prevalence of COVID-19 in CML patients, as of Feb 21, 2020. Secondary outcomes included the history of epidemiology of CML patients, the clinical characteristics and outcomes of CML patients with COVID-19.

### Statistical analysis

The aim of this study is to report prevalence of COVID-19 in CML patients. COVID-19 infection in CML patients with different efficacy was compared. We also report the clinical courses and clinical outcomes of the COVID-19 patients. There were, therefore, no formal hypotheses being implemented to drive the sample size calculation and we included the maximum number of patients who met the inclusion criteria.

## Results

A total of 413 questionnaires were collected, of which 392 were valid. Among them, 21 questionnaires were excluded because the respondents did not live in Hubei province during the outbreak. The following is the general situation of the respondents. There were 223 males, accounting for 56.9%, and 61.2% of the respondents were 50 years or younger. The age distribution chart is shown in Table 1. The other details are shown in Table 2. The epidemiological history is shown in Table 3. Specific data of clinical diagnosis and confirmed patients are shown in table 4.

**Table 1.**
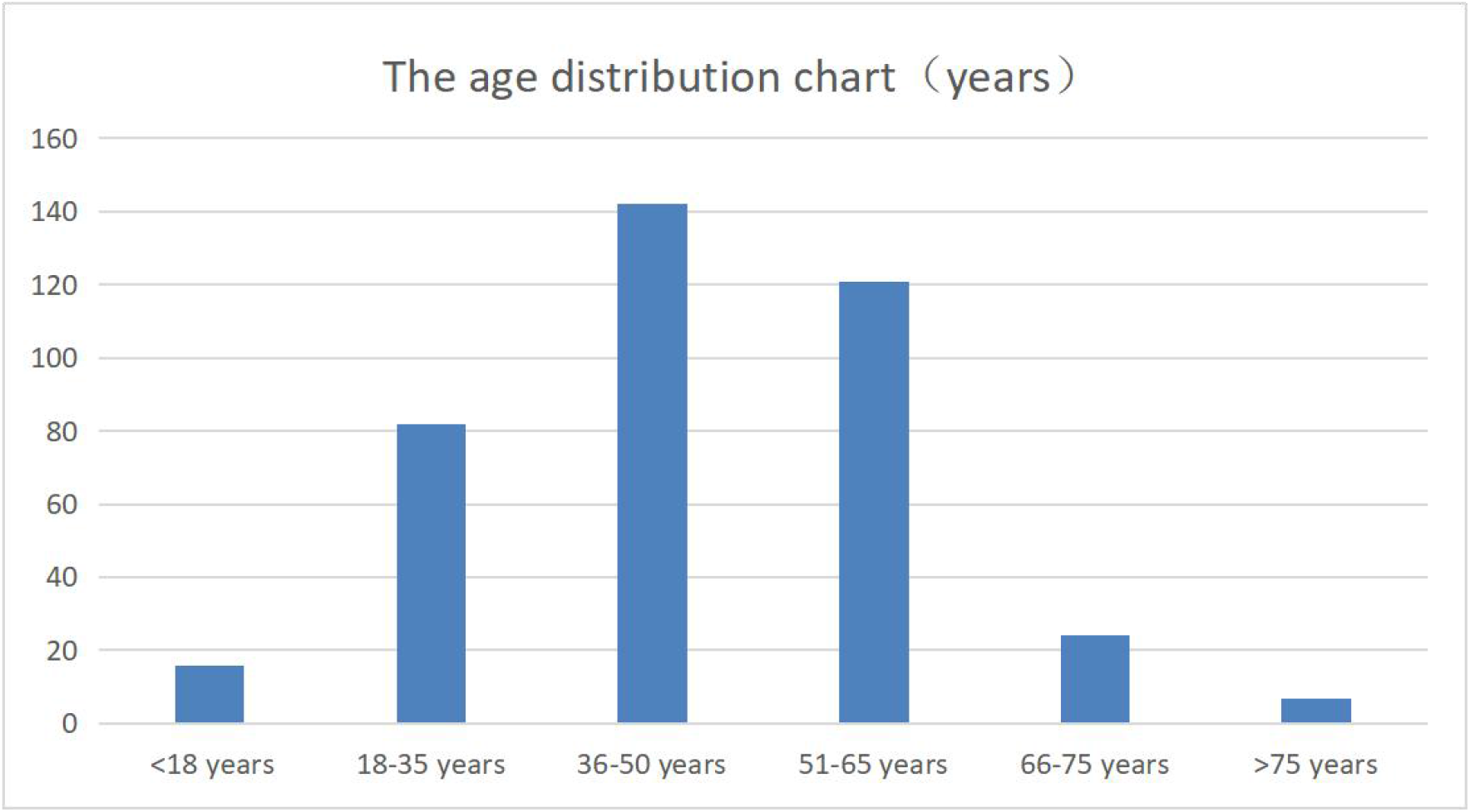
(the age distribution chart)

**Table 2.** (general demographic characteristics)

| General demographic characteristics | Count |
| --- | --- |
| Gender |  |
| Male | 223 |
| Female | 169 |
| Age(years) |  |
| Less than 18 years of age | 16 |
| 18-35 | 82 |
| 36-50 | 142 |
| 51-65 | 121 |
| 66-75 | 24 |
| More than 75 years old | 7 |
| Diagnose CML time |  |
| Less than 3 months | 9 |
| 3-12 months | 56 |
| 1-3 years | 136 |
| 3-10 years | 139 |
| More than 10 years | 52 |
| Current TKI status |  |
| Dasatinib | 82 |
| Imatinib | 259 |
| Nilotinib | 37 |
| Else | 14 |
| Efficacy evaluation of CML |  |
| Optimum response | 299 |
| Warnings | 20 |
| Failure | 30 |
| Not known | 43 |
| Comorbidities |  |
| Hypertension | 26 |
| Other Malignancies | 10 |
| Kidney diseases | 9 |
| Diabetes | 14 |
| Else | 53 |
| None | 280 |
| Combined medications |  |
| Corticosteroids | 10 |
| Antihypertensive drugs ( Angiotensin-Converting Enzyme Inhibitors ) | 32 |
| Other anti-tumor drugs | 3 |
| Else | 58 |
| None | 289 |
| Smoking history |  |
| Not have | 309 |
| Have | 83 |
| Have you forgotten to take medicine in the past 2 weeks |  |
| No | 315 |
| Yes | 77 ( 10 patients took drugs irregularly due to the influence of the epidemic ) |

**Table 3.** (the epidemiological histories)

| The epidemiological history | Count |
| --- | --- |
| Residence |  |
| Wuhan | 82 |
| Hubei province (except wuhan) | 310 |
| contact with infected person |  |
| Yes | 4 ( 3 were in remote contact and 1 was in close contact ) |
| No | 330 |
| not known | 58 |
| Cluster onset |  |
| yes | 1 ( 3 people confirmed of COVID-19 in her family. This respondent did not live with the COVID-19 patient, the respondent does not have an infection at present ) |
| no | 391 |

**Table 4.**
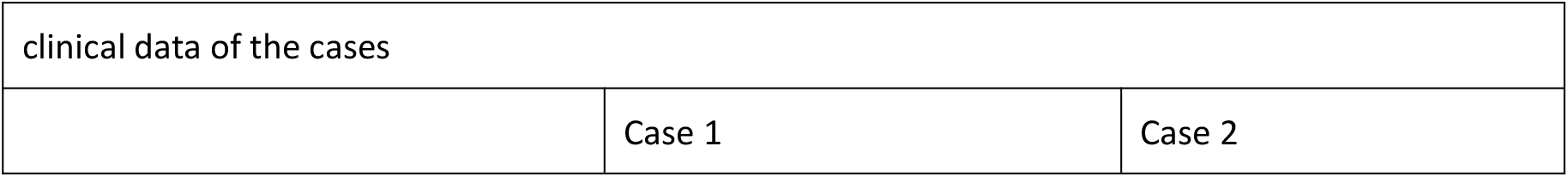
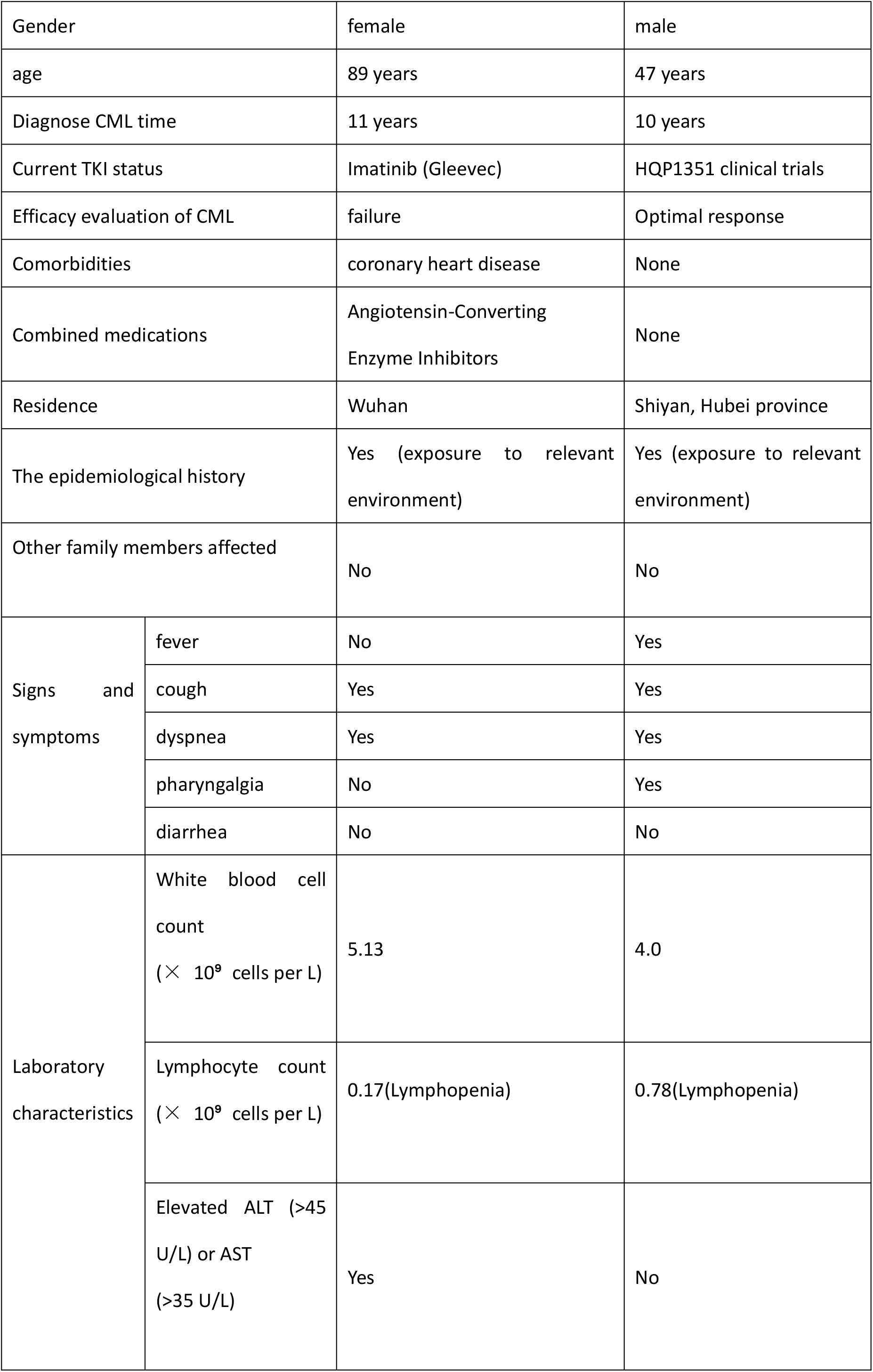
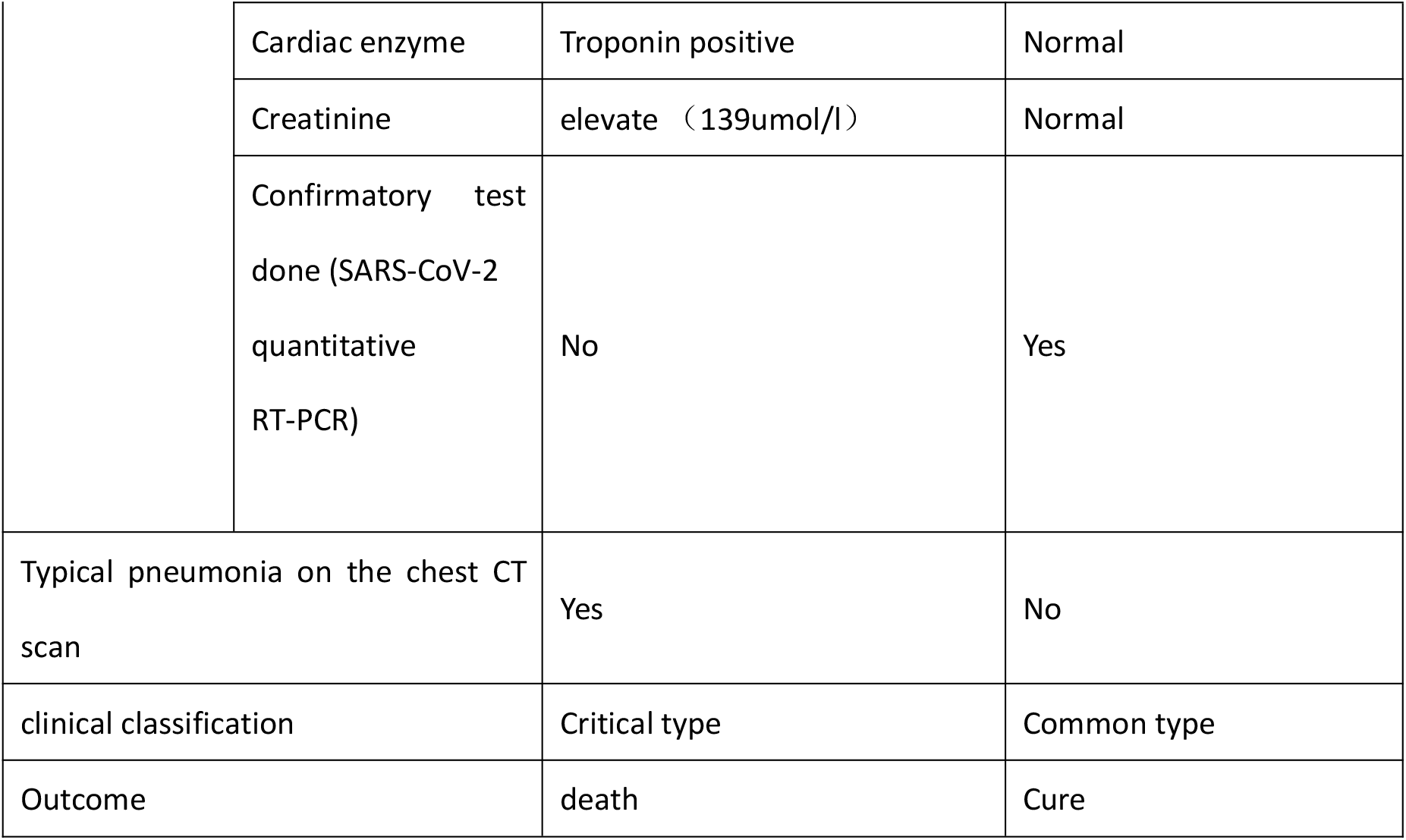
(clinical data of the cases)

The survey found that 12 respondents had fever, cough or shortness of breath during the epidemic period, and 9 of them did not go to the hospital because their symptoms were mild or remitted with oral medication. Three people went to the hospital for treatment. After chest CT scan and SARS-CoV-2 quantitative RT-PCR testing, one of these three patients was confirmed to have of COVID-19 infection and was treated and cured. COVID-19 infection was excluded in a second patient. The third patient was clinically diagnosed based on the 5th version of the novel coronavirus infection pneumonia diagnosis and treatment (National Health Commission of the People’s Republic of China) that was used at the time. This patient, who was clinically diagnosed and treated as a confirmed case, was an 89-year-old female with coronary heart disease and long-term ACEI treatment. She was diagnosed CML (Chronic phase) in January 2009. Before 2015, she did not take imatinib regularly due to side effects and the efficacy evaluation was failure. After 2015, regular dose reduction of imatinib (200mg per day) was started. The latest BCR-ABL fusion gene was quantified at 0.37% in December 2019. In early February 2020, after initially having a cough, she quickly developed dyspnea that was not relieved with oxygen therapy at home. She was admitted to the emergency department where she was found to have extreme lymphocytopenia, respiratory failure, renal insufficiency, and myocardial damage. Her condition rapidly deteriorated and she succumbed after three days of hospitalization. This is consistent with previous reports that elderly patients with comorbidities have a poor prognosis and high mortality^20-22^, and the degree of lymphopenia is related to the severity of the disease.

Another CML patient with a confirmed COVID-19 infection was a 47-year-old male. He was initially diagnosed with CML (accelerated phase) in January 2010. Treatment with imatinib was irregular after diagnosis. In 2015, dasatinib was replaced due to treatment failure. In March 2019, the efficacy evaluation was failure: the BCR-ABL fusion gene quantification was 25.47%, the chromosome was 46, XY, t (9;22) (q34;q11) [10] /47, XY, +8 [10] and there was a T315I mutation. He participated in an ongoing clinical trial of an alternative CML treatment (HQP1351) in May 2019 and achieved complete cytogenetic remission (CCyR) in July 2019. The chromosome was 47, XY, +8 [20], and the fusion gene quantification was 0.49%. Major molecular remission (MMR) was achieved in August 2019, and complete molecular remission (MR4.5) was achieved in October 2019. Although he obtained an optimal response, he was not taking TKIs during the outbreak due to side effects. In mid-february 2020, he presented with cough and fever and was found to be infected with SARS-CoV-2, but a chest CT scan did not show pneumonia and he was cured and discharged after treatment. The specific data of the clinical diagnosis and confirmed patients are shown in table 4.

## Discussion

While the 392 CML respondents required regular referrals to hospitals, they did not have much contact with COVID-19 patients during the outbreak, mainly due to the government policy of prevention and control.

We then analyzed SARS-CoV-2 infections in patients based on their response to anti-CML therapy (see Table 5 for details), and found that only 1 of 299 (0.3%) patients with an optimal response was diagnosed with COVID-19. Of the 50 patients who failed to respond to CML treatment or had a poor response, 1 patient (2%) had a clinical diagnosis of COVID-19. Thus patients who failed to achieved an optimal response to CML therapy appear more likely to have a symptomatic infection with SARS-CoV-2. This tendency requires further study, but there are two possibilities that might explain it. First, an optimal response to TKI treatment may be associated with immune recovery. CML patients exhibit selective depletion of effector T reg cells (eT reg cells) ^23,24^, while TKIs increase the number of natural killer cells (NK), NK-LGL and T-LGLs cells^25^, which play a role in regulating immunity. Second, previous studies have reported that imatinib and other TKI drugs have antiviral activity *in vitro* against Middle East respiratory syndrome coronavirus (MERS-COV) and severe acute respiratory syndrome coronavirus (SARS-COV)^26-28^. It will be interesting to see if TKIs also have activity against anti-SARS-CoV-2.

**Table 5.**
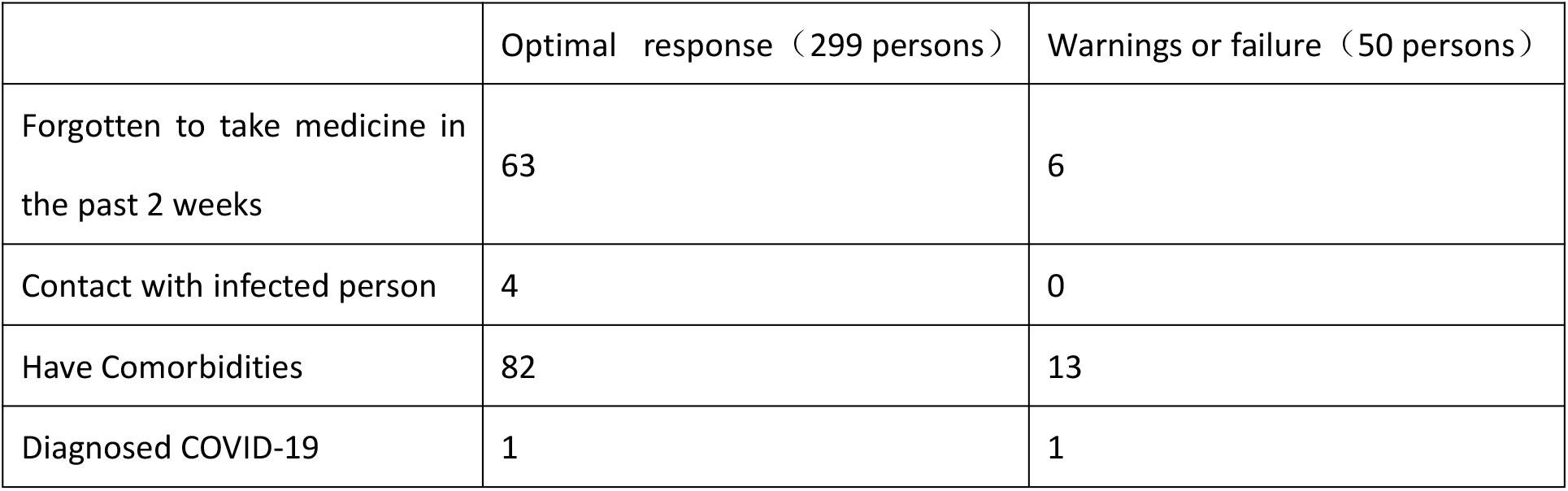
(Data were compared between the CML patients with optimal response and those with non-optimal response)

|  | Optimal response (299 persons) | Warnings or failure (50 persons) |
| --- | --- | --- |
| Forgotten to take medicine in the past 2 weeks | 63 | 6 |
| Contact with infected person | 4 | 0 |
| Have Comorbidities | 82 | 13 |
| Diagnosed COVID-19 | 1 | 1 |

We analyzed the 82 respondents living in Wuhan and found that there was 1 clinically diagnosed case (1.2%). Of the Japanese citizens evacuated from Wuhan, 8/565 (1.4%) had a confirmed infection with COVID-19 ^29^. Among 310 CML living patients in Hubei province outside of Wuhan, 1 case of infection (0.32%) was diagnosed.

In summary, through the Hubei Anti-Cancer Association Chronic Myeloid Leukemia Standardized Management Collaboration Group, we conducted online consultations, online smart hospitals and online courses to provide information, diagnosis and treatment to patients during the epidemic, and organized volunteers to deliver drugs to CML patients at their homes. The sample size of the study was relatively small and should be expanded to more accurately compare the prevalence of the infection in the different groups. Nevertheless, our findings suggest that standardized management to obtain the best clinical response reduces the risk of SARS-CoV-2 infection and improves the prognosis of those who are infected. CML is a chronic hematologic tumor, and comprehensive measures should be taken to delay and reduce the risk of infection by restricting patient exposure to situations with possible transmission. During the epidemic, special attention should be focused on those CML patients who are older, have co-morbidities and whose response to CML treatment is non-optimal.

## Data Availability

All data generated or analyzed during this study are included in this artical.

